# VaxiMap: optimal delivery of vaccinations for housebound patients

**DOI:** 10.1101/2021.12.20.21267978

**Authors:** Thomas F. Kirk, Adam J. Barker, Armen Bodossian, Robert Staruch

## Abstract

**Background:** Throughout the UK’s Covid-19 vaccination campaign, responsibility for vaccinating housebound patients has rested with individual GP surgeries, posing them a difficult logistical challenge (the travelling salesman problem). In response to demand from GPs, and a lack of existing solutions tailored specifically to vaccination, VaxiMap was created. This tool provides optimal routes for vaccine delivery and has been free to all users since its inception in January 2021.

**Methods:** VaxiMap generates optimal routes subject to the constraint that the number of patients per route should be fixed. This ensures that a known quantity of vaccine can be set aside for each route and minimises wastage. The user need only upload an Excel spreadsheet of patient postcodes to be visited. A divide-and-conquer approach of iterative *k*-means clustering followed by within-cluster route optimisation is used to generate the routes.

**Findings:** We find substantial savings in the time taken to plan vaccinations, as well as savings in the time taken to visit housebound patients. We estimate total savings to date of 4,700 hours of practitioner time, equivalent to 2.5 work-years, or approximately £91k at typical practitioner salaries.

**Interpretation:** The adoption of VaxiMap yielded both time and cost savings for GP surgeries and accelerated the UK’s Covid-19 vaccination campaign at a critical moment.

**Funding:** Financial support for VaxiMap was provided by Magdalen College, Oxford, Oxford University Innovation, and JHubMed, part of UK Strategic Command. These parties were not involved in the preparation of this manuscript.

## 1. Background

In early 2021 a global campaign to vaccinate against SARS-COV2 was launched. In the UK, the Joint Committee on Vaccination and Immunisation (JCVI) stipulated that patients should be vaccinated in descending age and risk order [1]. Under the nine risk categories identified by the JCVI, housebound patients were allocated into the higher priority groups for early vaccination.

There are estimated to be on the order of half a million housebound patients in the UK^1^. Responsibility for vaccinating these patients has rested with general practitioner (GP) surgeries as they cannot attend a mass vaccination centre. In order for the wider vaccination campaign to proceed on schedule, it was therefore of vital importance that GPs could quickly and efficiently deliver vaccinations to these patients. Given their high risk profile, timely vaccination would also reduce the burden on other parts of the health system. The emergence of new variants suggests that mass vaccination will continue to be required for some time to come in order to keep the pandemic under control.

The central problem in optimising the vaccination of housebound patients is determining the fastest order in which to visit the individuals. This is an example of the travelling salesman problem (TSP), which has been extensively studied in the domain of computer science. Notwithstanding numerous solutions to the problem [2], some of which have been applied in a medical context [3], vaccination imposes an extra constraint: because there are a fixed number of doses in a vial, it is preferable sort patients into groups of this same number. This ensures that exactly one vial (or integer multiples thereof) of vaccine will be required per group of visited patients. Separately, due to the cold-storage requirements of the vaccines themselves, visiting patients in the fastest order minimises the time that vials spend outside of the cold chain. Taken together, these two factors reduce vaccine wastage, especially important given the limited availability of supplies during the early stages of the campaign.

Though commercial solutions for route planning exist, these were not widely adopted by GPs for use in the vaccination campaign, possibly because they do not allow for constraints on group size. In response to numerous requests for help on social media from GPs and practice managers, it was therefore decided to build a domain-specific solution entitled VaxiMap (www.vaximap.org) in January 2021. The service is a free-to-use website that requires only an Excel spreadsheet of patient postcodes to function. The patients are sorted into groups of fixed size based on proximity to each other, the fastest route within each group found, and directions and estimated travel times returned to the user (an example is shown in figure 1). Within a month of operation, 100,000 vaccine deliveries had been planned on the site and the service was adopted for use by military vaccination quick reaction force teams during operation RESCRIPT, the UK military’s support to HM Government during the pandemic. As of December 2021, over 350,000 deliveries have been planned.

**Figure 1:**
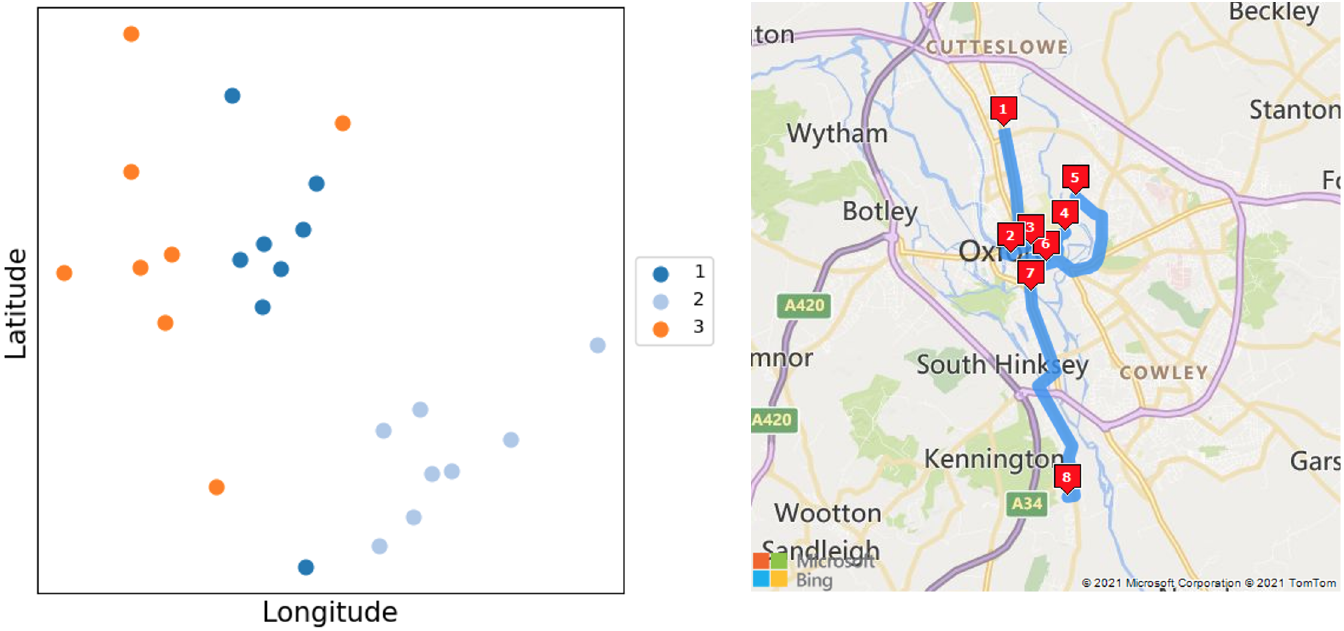
Example output produced by VaxiMap. Left: 24 patients have been sorted into three clusters of size eight, the spatial distribution of which is shown here. Right: the optimal driving route for visiting the patients in cluster no. 2 (dark blue).

The purpose of this paper is to explain how the service functions and to estimate its beneficial impact to date (via time savings in vaccine delivery). The data findings presented are of interest to public health officials involved in the delivery of services to housebound patients, be they vaccination-related or otherwise.

## 2. Methods & Materials

### 2.1. Implementation

The problem is posed as finding the shortest routes to visit a set of *N* patients, whilst ensuring any individual practitioner visits no more than *D* patients per route. It is assumed, but not required, that *D* is set as the number of vaccine doses in a vial (for example, nine for Oxford-AstraZeneca); any number between 3 and 25 can be used. It follows that the *N* patients must be sorted into *G* = ceiling(*N/D*) groups (rounded up in the case that *D* does not divide perfectly into *N*, in which case *exactly* one group will have size less than *D*)^2^.

Patient postcodes uploaded by the user are transformed into latitude and longitude coordinates on the *x, y* plane via the use of Microsoft Bing’s geocoding service. Next, the patients are grouped so that they are proximal in space, a simple heuristic to minimise the travel time *within* each group. Iterative *k*-means clustering is used to group the patients subject to the constraint on group size *D*, which standard *k*-means cannot do [4, 5]. Qualitatively, this approach assigns those patients that are furthest away from all others first, as these must be placed into the ‘correct’ group, whereas those that are close to the centre of the distribution can be assigned to any group with little consequence. The clustering process is illustrated in figure 2.

**Figure 2:**
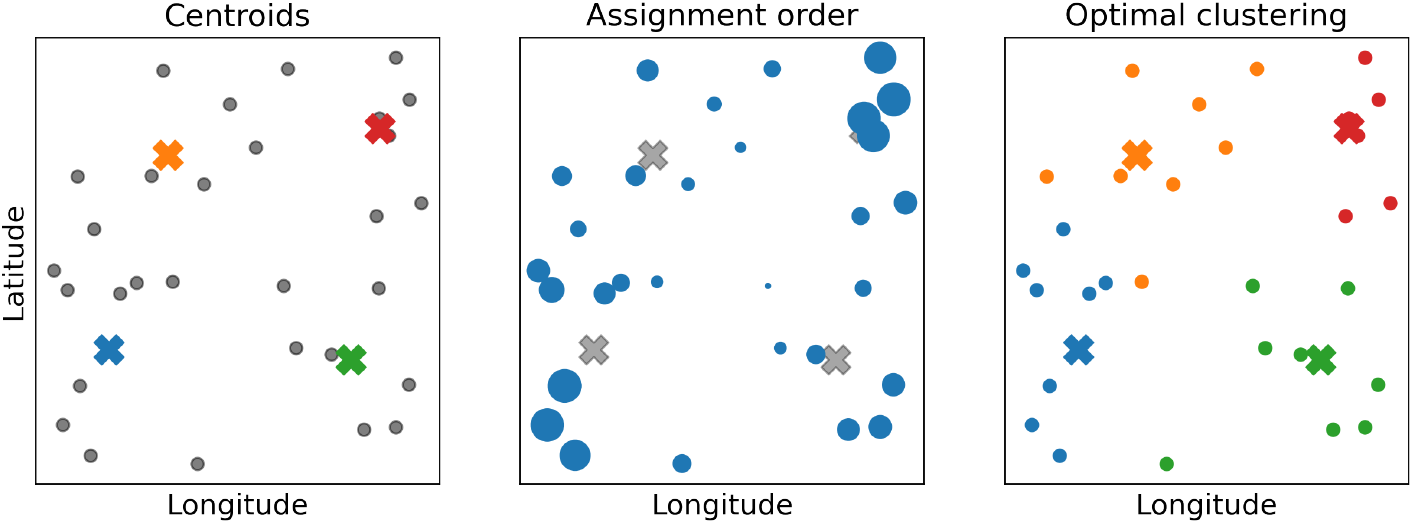
Example clustering of *N* = 30 patients into groups of *D* = 8. Left: initialisation of four cluster centroids (denoted with crosses). Centre: order of patient assignment, where larger circles indicate priority. Patients in the centre of the distribution could be assigned to any cluster with low cost, so are dealt with last. Right: the optimal clustering of patients after assignment. The red cluster has six patients due to imperfect number division, whereas all other clusters are fully-sized.

After cluster assignment, the final step is to determine the optimal order in which to visit the patients of each group. This is achieved via Microsoft Bing’s mapping API, making use of the *optimise waypoints* facility (which imposes a limit of 25 patients per group) [6]. Optionally, the user can specify a fixed start and end point for their routes (for example, the address of their GP surgery); each route will then be a round-trip from that location. The user is returned the patient groups, the optimal driving or walking directions within each group, and travel time estimates for each group.

### 2.2. Dataset

Since the launch of the VaxiMap service in January 2021, a database of generated routes has been accumulated (a consent statement is given in the supplementary material). From each user *request* (corresponding to a single upload of patient postcodes and the routes that result), the following data are retained: time and date; number of patients; requested cluster size; relative distances between patients (not absolute locations); and transport mode. For approximately a third of requests, counts of patient postcode districts^3^ were also retained (postal district is the first part of a postcode, for example OX1). As of December 2021, the dataset comprises 11,000 requests for 350,000 patients; postal district information is available for 4,700 of the requests.

### 2.3. Analysis methods

The objective of analysis was to estimate the time savings yielded by VaxiMap and to identify high-level trends in how the service has been used. The savings arise in two ways: firstly, when *planning* a route to visit patients; and secondly when *following* an optimal route instead of a sub-optimal one. The following analyses were performed.

#### Repeat detection

In the course of development, it was noted that users sometimes uploaded the same set of patients multiple times in quick succession, with slightly different parameters. It is suspected that such behaviour reflects a learning process on the part of users who were familiarising themselves with the site. For certain analyses detailed below, such repeat requests (defined by creation within 21 days of a previous identical request) were removed. It was not desired to remove repeats separated by more than 21 days as these could represent genuine repeat uses, though they may not correspond to repeat vaccinations if separated by just a few weeks.

#### Summary metrics

Summary metrics of the dataset were explored by drawing histograms of the number of patients uploaded per request, the number of clusters the request was split into, and the number of patients per cluster. The time-series nature of the data was explored by plotting the number of user requests per day, and the total number of patients across all requests per day. Finally, the geographic distribution of the data was exploited by plotting the cumulative number of patients across all requests in each UK postal district for the subset of data with this information. These analyses were performed on the dataset including repeats.

#### Route planning

The time taken for humans to plan solutions for the TSP is remarkably quick and scales linearly with the number of locations to be visited [8, 9, 10]. The problem solved by VaxiMap is subtly different to the conventional TSP investigated in the literature because users start with a text-based list of addresses (*i.e*, from an EMIS database) as opposed to a visual representation. This difference is important in light of the consensus view that “humans require a visual representation of the problem” in order to solve it effectively [10]. The time taken to plan routes in the absence of VaxiMap can therefore be split into two components: a *lookup time* to generate a spatial representation of the problem, and a *routing time* to actually plan the routes using this representation. A survey that investigated these components separately was conducted across 20 volunteers (8 female, mean age 45 years, further information given in the supplementary material). Linear regressions across their responses yielded the following estimates for the time taken to perform these tasks manually: 36·4 seconds/location for lookup time, and 4·8 seconds/location for planning time (uncertainties are quantified in the supplementary material). These coefficients were then multiplied across the entire dataset, including repeats, to obtain estimates of the time saved in planning. Repeats were included in this analysis as it was assumed users had reasonable cause for them (for example, experimenting with different cluster sizes and observing the differing travel times that result before making a decision).

#### Route following

The quality of human solutions to the TSP has been investigated extensively [8, 10, 11, 12]. These are often remarkably close to optimal for small problems and scale well for larger problems (*n >* 50 locations). The empirical model of performance given in figure 2b of Dry’s review [9], reproduced below in figure 3, was used in this work to approximate the extra distance penalty *p*(*n*) of human solutions compared to VaxiMap’s solution. The penalty is very small for problems with *n ≤* 25 patients. For each user request in the non-repeated dataset, the total (closed) length of the VaxiMap generated routes *L*_*vm*_ was calculated using the method given in the supplementary material. The approximate length of the human solution *L*_*h*_ to the same problem was then obtained by multiplication with a scaling factor of (1 + *p*(*n*)) drawn from figure 3. In order to convert distance savings into time savings, a mean driving speed of 50 km/h or 30 mph was assumed [13]. Repeats were not included in this estimate as it was assumed to be unlikely that the journeys themselves were undertaken (though a user may have planned the same route twice in a week, they are unlikely to have actually undertaken it twice).

**Figure 3:**
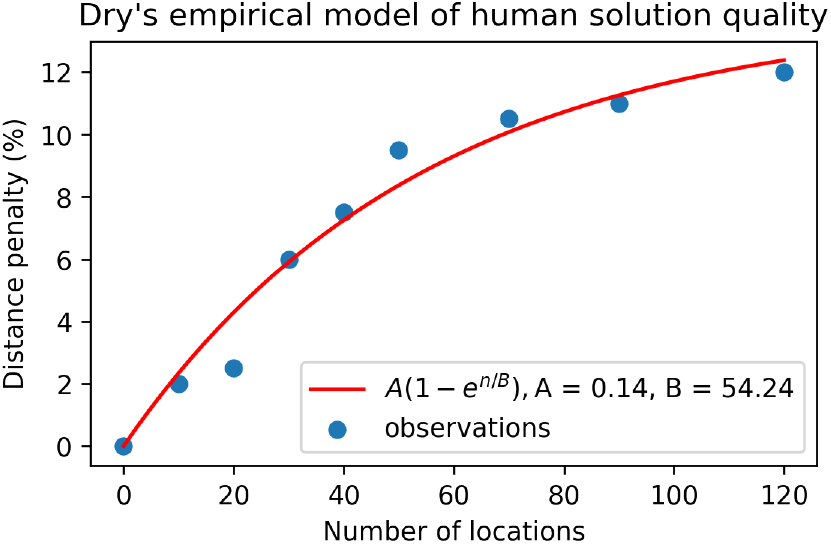
An empirical model of human performance on the TSP, taken from Dry’s review [9]. The individual observations from that work are reproduced here; a curve fit of the form *A*(1*−e*^*n/B*^) was performed to yield a smooth model. The penalty is less than 10% for problems sized up to around 60 locations.

### 2.5. Ethics statement

This work has been classified as service development and evaluation by the Medical Sciences Interdivisional Research Ethics Committee at the University of Oxford, and does not therefore require ethical review (CUREC application R79436/RE001).

## 3. Results

### 3.1. Summary metrics

Figure 4 shows the timeseries distribution of VaxiMap use, both in terms of total number of daily patients, and patients per user request. Three peaks that are assumed to correspond to first, second and booster doses can be observed.

**Figure 4:**
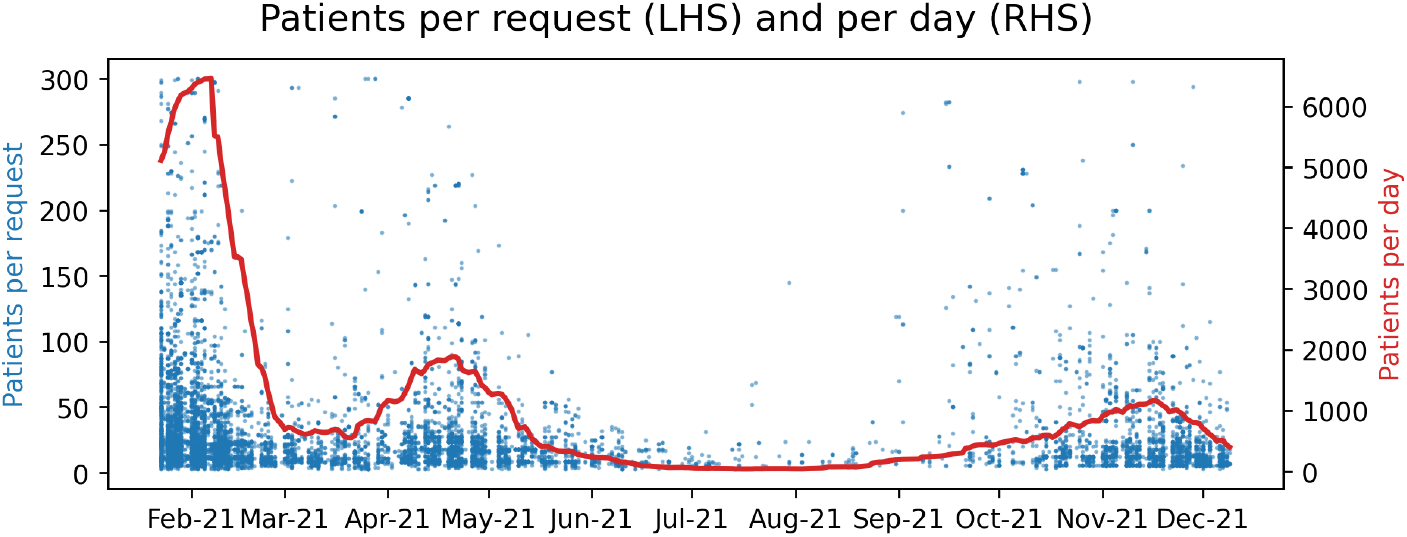
Timeseries of VaxiMap use. Peaks assumed to correspond to the majority of first (Jan ‘21) and second doses (Apr ‘21) can be discerned, as can booster doses (Oct ‘21). The red line is the 30-day moving average of total uploaded patients per day. A few uploads of the maximum permitted of 300 patients can be observed throughout the period.

Figure 5 shows the geographic distribution of uploaded patient locations within the UK, for a subset of the complete dataset. Use of the service has been concentrated in England and Wales; by contrast Scotland and Northern Ireland have seen very little use.

**Figure 5:**
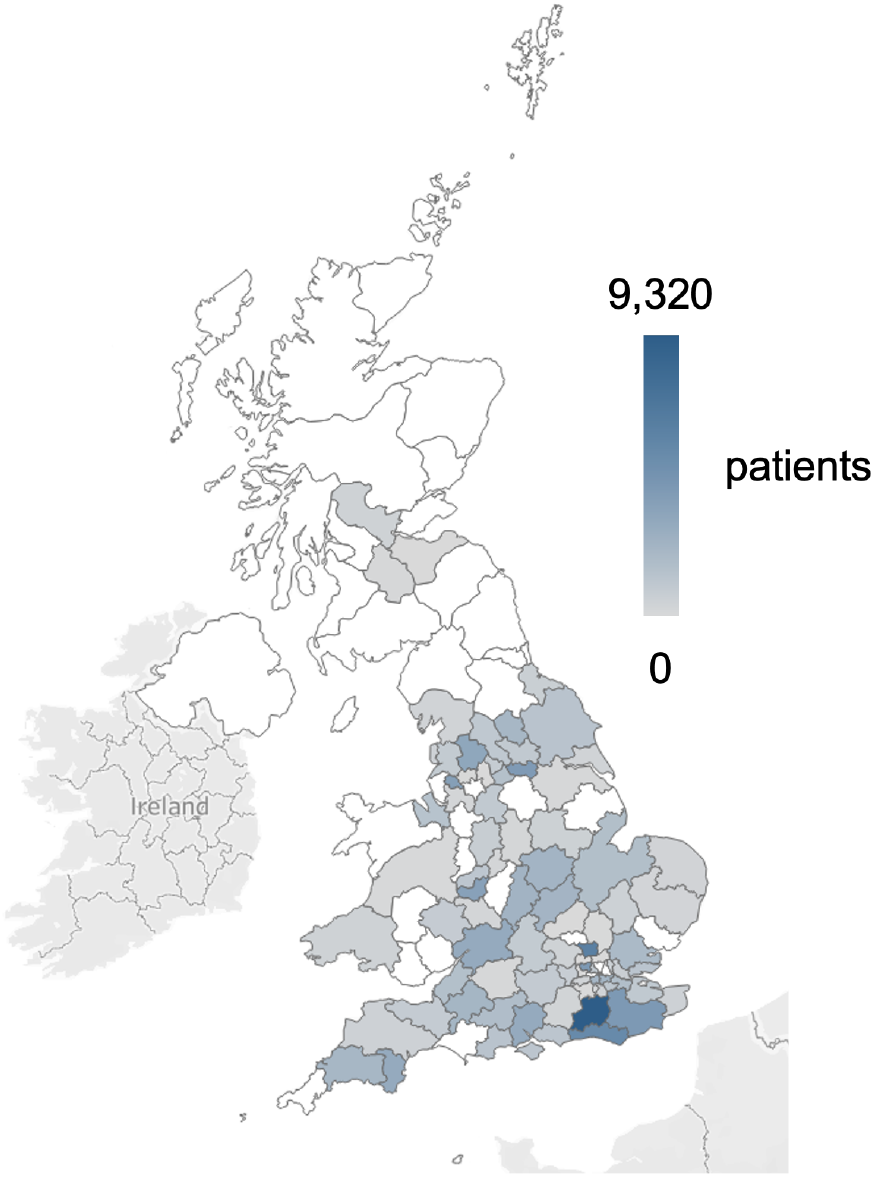
Distribution of processed patient locations within UK postal districts, for a subset of the complete datset. White denotes no data.

Figure 6 shows summary statistics of uploaded user requests. Users uploaded a median of 18 and mean of 32 patients per request, with a target cluster size of around 10. Though a handful of users uploaded the maximum limit of 300 patients, a substantial minority uploaded 10 or so patients, resulting in a single cluster being generated.

**Figure 6:**
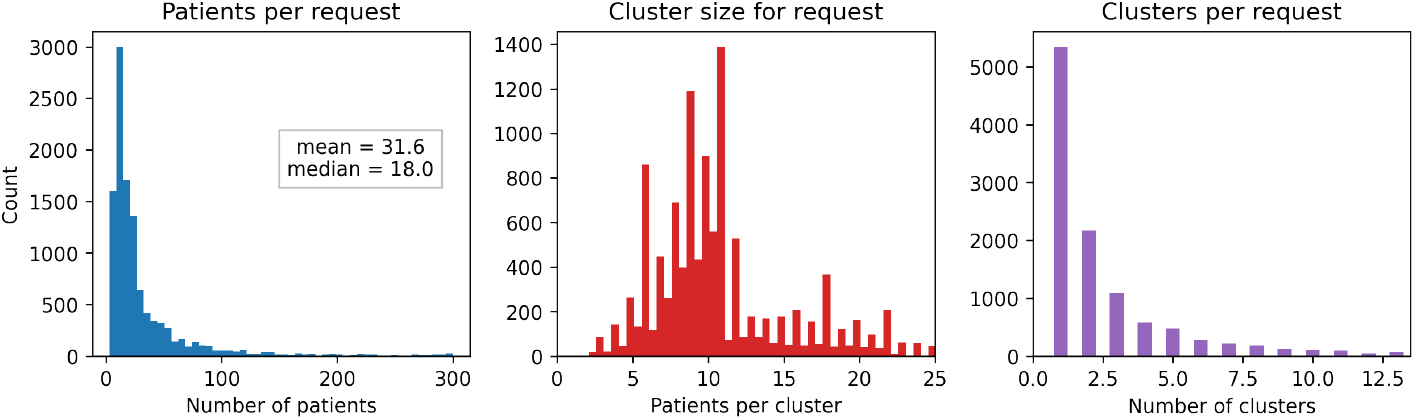
Histograms of user request characteristics. Left: total number of patients uploaded. Centre: request cluster size. Right: number of clusters per request.

### 3.2. Repeat detection

Using a 21 day repeat threshold, 3,391 repeat requests were detected. The dataset excluding 21-day repeats was thus reduced from 11,105 requests covering 351,319 patients to 7,732 requests covering 224,692 patients. The latter figure represents the best estimate of the number of home visits that have been performed using the service. Of the repeats that remained within the dataset (separated by more than 21 days), some, but not all, could be consistent with second or booster vaccination doses. An example of a repeat that cannot be explained by vaccination is shown in figure 7: there are four occurrences across five months, all separated by at least one month. Further examples of repeats are given in the supplementary material.

**Figure 7:**
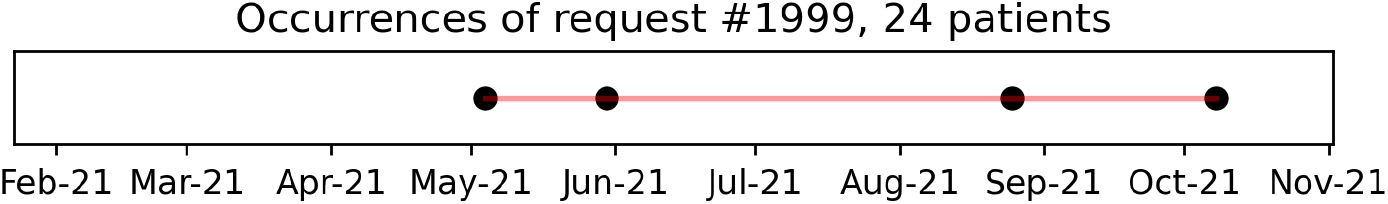
A single example of repeat usage that is inconsistent with vaccination: there are too many occurrences (four) over too short a time period (five months).

### 3.3. Route planning

Applied to the full dataset including repeats, the survey-derived lookup times and routing times of 36·4s and 4·8s per location, respectively, yielded an estimate of total time savings in planning of 4,020 hours, equivalent to 100 weeks at 40 hours/week.

### 3.4. Route following

Applied to the full dataset without repeats, an empirical model of human performance on the TSP yielded an estimate of the total savings in distance travelled of 35,184 km. This is equivalent to 703 hours, or 17 weeks at 40 hours/week, assuming a mean travel speed of 50 km/h or 30 mph.

Combining all savings together yielded a total of 4,724 hours of practitioner time, equivalent to 118 work-weeks or almost 2.5 work-years. The time savings break down approximately in a 4:1 ratio for planning to travelling. Using mean salary estimates of £38k and £32k for a practice manager and community nurse respectively^4^, these time savings can be converted into a cost saving of approximately £91k.

## 4. Discussion

We have presented a simple and easy-to-use solution for optimising vaccine delivery to housebound patients. It has seen widespread use during the UK’s Covid-19 vaccination campaign, reaching close to 50% of the target patient population^5^, and yielded both time (over two years of practitioner time) and cost (approximately £91k) savings. One user in Plymouth reported doubling their rate of delivery through using the service, a finding that implies VaxiMap was able to reduce the burden on other parts of the healthcare system by quickly removing vulnerable patients from the unvaccinated population.

Harder to quantify are the savings in cognitive load of automating the complicated process of route planning, but direct feedback from users (published on the VaxiMap website) frequently touched on the ‘frustration’ inherent to such a tedious and labour-intensive task. This is especially relevant given the small number of users that uploaded requests containing the maximum allowed number of 300 patients; a manual approach for this number would be infeasible. In light of the very low investment that was required to set up this service (the beta version was online in 48 hours), the overall cost-benefit ratio of this simple intervention is extremely favourable.

The development of VaxiMap relied upon real-time feedback from users. For example, the addition of walking directions (alongside driving) was made in response to requests from GP surgeries in urban areas. Widespread adoption was achieved almost entirely through word of mouth (in particular, GP networks on social media) with no engagement from the NHS itself. This shows the importance of interacting directly with users before and during development to ensure their requirements are met, which is pertinent in light of the expectation that digital technologies will play an ever-greater role in primary care [14].

At the time of writing, the service is supporting the booster campaign. However, the underlying problem that the service addresses will continue to exist long after Covid-19; namely, how to efficiently visit a set of patients subject to some constraint on group size? The underlying technology could therefore be applied in other domains. An obvious example, and one already suggested by multiple users, would be supporting annual flu vaccination campaigns; a more novel example would be supporting district nurses, community nurses and physiotherapists in their day-to-day duties. Intriguingly, the analysis of repeat requests reveals use patterns that are inconsistent with vaccination, which suggests that some users have already employed the service for other purposes. Given that community healthcare in the UK records around 100 million patient contacts annually with a budget of around £10 billion and one-fifth of the NHS workforce [15], the cumulative impact of efficiencies obtained at the grassroots level could be substantial.

## Data Availability

All data produced in the present study are available upon reasonable request to the authors

## 5. Contributions

TFK developed the VaxiMap software and website, and set up data collection. RS identified the core need, found a community of beta testers, and acted as clinical liaison for users throughout. AJB and AB performed survey and data analysis. All authors prepared and approved the manuscript. TFK, AJB and AB verified the data. All authors had access to the data and accompanying analysis. AJB, AB and RS were acting independently of their respective employers; this work has not been endorsed by, nor does it reflect the views of, Squarepoint Capital LLP, Vodafone Group PLC, Defence Medical Services or the Ministry of Defence.

## 7. Acknowledgements

The authors acknowledge financial support provided by Magdalen College, Oxford, Oxford University Innovation, and JHubMed, part of UK Strategic Command. Support of a technical nature was provided by Microsoft Bing. The authors thank Dr David Andrews for valuable feedback during the development of the service, and Captain Amrit Sandhu for integrating support from JHubMed.

## 8. Data sharing

The authors intend to make the VaxiMap dataset freely available following peer review. This will be via insertion into the Oxford Research Archive with accompanying DOI. The source code used for analysis of the dataset will also be released on GitHub. Until such a time, readers should contact the corresponding author for all enquiries.

## Footnotes

1 Numerous freedom of information requests to NHS England, Public Health England and the JCVI have not yielded a precise figure. Correspondence with a member of the Health Informatics Group at the Royal College of General Practitioners suggests a lower bound of 250,000.

2 This is an important extra constraint added at the request of a GP, which ensures at most one vial of vaccine will be incompletely consumed.

3 Typical postal districts have in excess of 10,000 patients [7], so this information is geographically non-precise.

4 Taken from www.glassdoor.com

5 Assuming a patient population of 500,000 and using a figure of 225,000 non-repeated visits planned on the site.

## References

[1] Joint Committee on Vaccination and Immunisation. Advice on priority groups for COVID-19 vaccination, 30 December 2020. Technical report, Department of Health and Social Care, 2020. URL https://www.gov.uk/government/publications/priority-groups-for-coronavirus-covid-19-vaccination-advice-from-the-jcvi-30-december-2020/joint-committee-on-vaccination-and-immunisation-advice-on-priority-groups-for-covid-19-vaccination-30-december-2020.

[2] Gilbert Laporte and Silvano Martello. The selective travelling salesman problem. Discrete Applied Mathematics, 26(2):193–207, 1990. ISSN 0166-218X. doi: https://doi.org/10.1016/0166-218X(90)90100-Q. URL https://www.sciencedirect.com/science/article/pii/0166218X9090100Q.

[3] Yufen Shao, Jonathan F. Bard, and Ahmad I. Jarrah. The therapist routing and scheduling problem. IIE Transactions (Institute of Industrial Engineers), 44(10):868–893, 2012. ISSN 0740817X. doi: 10.1080/0740817X.2012.665202.

[4] James MacQueen and Others. Some methods for classification and analysis of multivariate observations. In Proceedings of the fifth Berkeley symposium on mathematical statistics and probability, number 14, pages 281–297. Oakland, CA, USA, 1967.

[5] Ian Davidson and S S Ravi. Clustering with constraints: Feasibility issues and the k-means algorithm. In Proceedings of the 2005 SIAM international conference on data mining, pages 138–149. SIAM, 2005.

[6] Microsoft. Bing Maps Routes API, 2020. URL https://docs.microsoft.com/en-us/bingmaps/rest-services/routes/.

[7] Office for National Statistics. Population for postcode districts in England and Wales, 2015. URL https://www.ons.gov.uk/aboutus/transparencyandgovernance/freedomofinformationfoi/populationforpostcodedistrictsinenglandandwales.

[8] J. N. Macgregor and T. Ormerod. Human performance on the traveling salesman problem. Perception and Psychophysics, 58(4):527–539, 1996. ISSN 00315117. doi: 10.3758/BF03213088.

[9] Matthew Dry, Michael D. Lee, Douglas Vickers, and Peter Hughes. Human Performance on Visually Presented Traveling Salesperson Problems with Varying Numbers of Nodes. The Journal of Problem Solving, 1(1):20–32, 2006. ISSN 1932-6246. doi: 10.7771/1932-6246.1004.

[10] James N. MacGregor and Yun Chu. Human Performance on the Traveling Salesman and Related Problems: A Review. The Journal of Problem Solving, 3(2):1–29, 2011. ISSN 1932-6246. doi: 10.7771/1932-6246.1090.

[11] Douglas Vickers, Marcus Butavicius, Michael Lee, and Andrei Medvedev. Human performance on visually presented Traveling Salesman problems. Psychological Research, 65(1):34–45, 2001. ISSN 03400727. doi: 10.1007/s004260000031.

[12] James N. MacGregor, Thomas C. Ormerod, and Edward P. Chronicle. Spatial and contextual factors in human performance on the travelling salesperson problem. Perception, 28(11):1417–1427, 1999. ISSN 03010066. doi: 10.1068/p2863.

[13] Praekash Balendra. Vehicle Speed Compliance Statistics, Great Britain: January - June 2020. Technical Report September, Department for Transport, 2020. URL https://www.gov.uk/government/statistics/vehicle-speed-compliance-statistics-for-great-britain-january-to-june-2020.

[14] World Health Organization (WHO). Digital technologies: shaping the future of primary health care. page 12, 2018. URL https://apps.who.int/iris/bitstream/handle/10665/326573/WHO-HIS-SDS-2018.55-eng.pdf.

[15] The King’s Fund. Community health services explained, 2019. URL https://www.kingsfund.org.uk/publications/community-health-services-explained.

