## Supplementary material for "VaxiMap: optimal delivery of vaccinations for housebound patients"

### Vaximap: optimal delivery of vaccinations for housebound patients
*supplementary material*

---

---

#### 1. Data collection and consent

The VaxiMap dataset was collected in accordance with the service's privacy policy and terms of use (https://vaximap.org/policy/). All users of the site consent to data collection subject to these terms. The data collected by the site is non-identifying: patient-specific postcodes are not retained, only postal districts. To each set of patient locations, a distance-preserving and non-reversible transformation that shifts their coordinates to the origin (0,0) of the global latitude and longitude coordinate system is applied. This means it is impossible to recover their original location.

#### 2. Survey methods

*Cohort.* There were 20 respondents, 8 female, with a mean age of 45 years ($\sigma$ = 19, minimum 26, maximum 78). Six respondents had occupations that are explicitly scientific, computational or data-focussed in nature.

*Questions.* Three questions investigated lookup time: respondents were presented with a list of postcodes of lengths 5, 8 and 13, respectively, and asked to locate them on map. They were allowed to use any tool to assist them in this, digital or otherwise, and were asked to record the time taken to complete each question. Three questions investigated routing time: respondents were presented with scatter-plot distributions of points, sized 13, 20 and 26, respectively, and asked to generate routes with a target size of 7 to visit all locations in as short a distance as possible. Again they were asked to record the time taken.

*Analysis.* In keeping with previous findings that human solution times scale linearly with problem size, linear regressions were performed on the survey responses to obtain estimates of lookup and planning time per patient. To each survey response, an extra datapoint of (0,0) was added to reflect the fact solution time should be zero when there are no patients. For both regressions, two extremely high survey response times were excluded as outliers, though they are presented in the results for completeness.

*Results.* Figure 1 shows the time taken to generate a spatial representation of a TSP as a function of the number of locations to be visited. A central estimate of 36·4s per location was derived from the regression ($R = 0{\cdot}78$). Figure 2 shows the time taken to generate a set of routes from a spatial representation of a TSP as a function of the number of locations to be visited. A central estimate of 4·8s per location was derived from the regression ($R = 0{\cdot}76$).

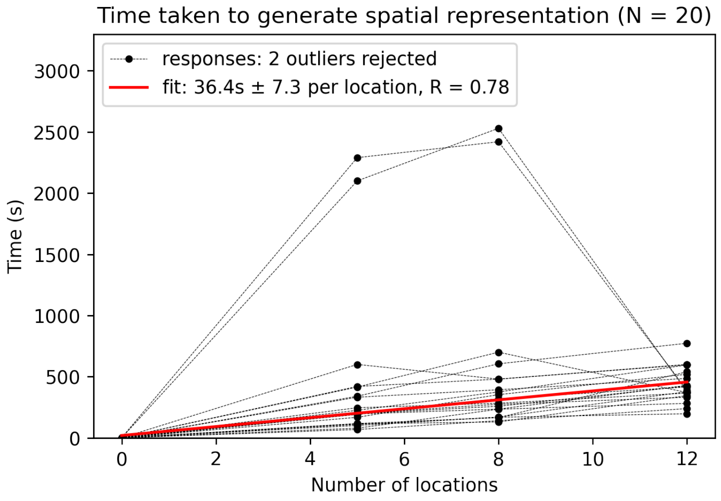


Figure 1: Time taken to locate patients on a map, expressed as a midpoint in a 95% confidence interval. Two survey responses were deemed outliers and excluded from the regression. The two extremely high response times came from respondents who used a physical map to complete the task (as opposed to a digital service such as Google Maps).

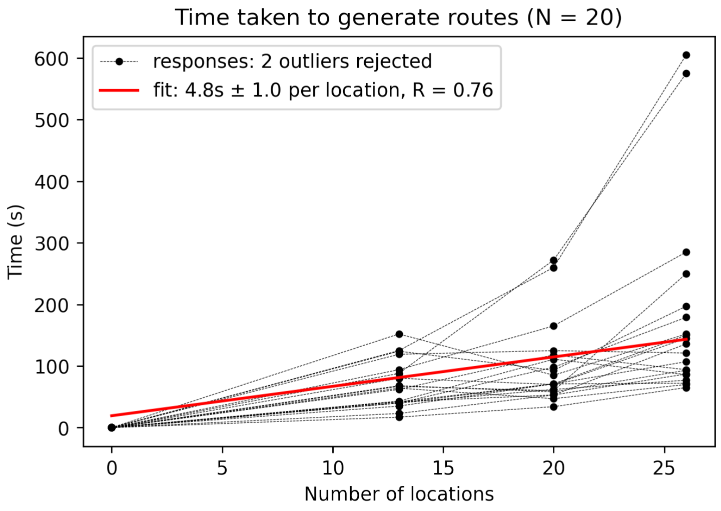


Figure 2: Time taken to generate routes on a map, expressed as a midpoint in a 95% confidence interval. Two survey responses were deemed outliers and excluded from the regression.

#### 3. Calculation of VaxiMap route lengths

The VaxiMap dataset contains the relative latitude and longitude coordinates for each set of uploaded patients. These have been shifted such that the centroid of each set lies on the origin (0,0), a non-reversible transformation that obscures the true location of the patients. The clustering and optimal orders in which to visit the patients are also recorded.

In order to calculate the length of routes between patients, it is necessary to calculate the pair-wise distances between patients. To do so, the patients are first shifted onto the UK by adding (53, -1.2), the approximate mid-point of the UK, to the coordinates. The coordinates are then transformed from latitude and longitude into a Cartesian metric coordinate system (EPSG:3857). From this, all pair-wise distances can be found by calculating the distance matrix for the patients, and finally closed route lengths are found by summing individual pair-wise distances as dictated by the optimal route orders.

In the real world, the shortest possible path between two points is rarely straight. The calculation performed above will give an artificially small estimate of total route length by assuming the existence of straight line paths between all patients. In order to correct this discrepancy, a multiplication by a *detour index* (ratio of real world distance to straight line distance) of 1·4 was performed [1].

#### 4. Repeat analysis: results

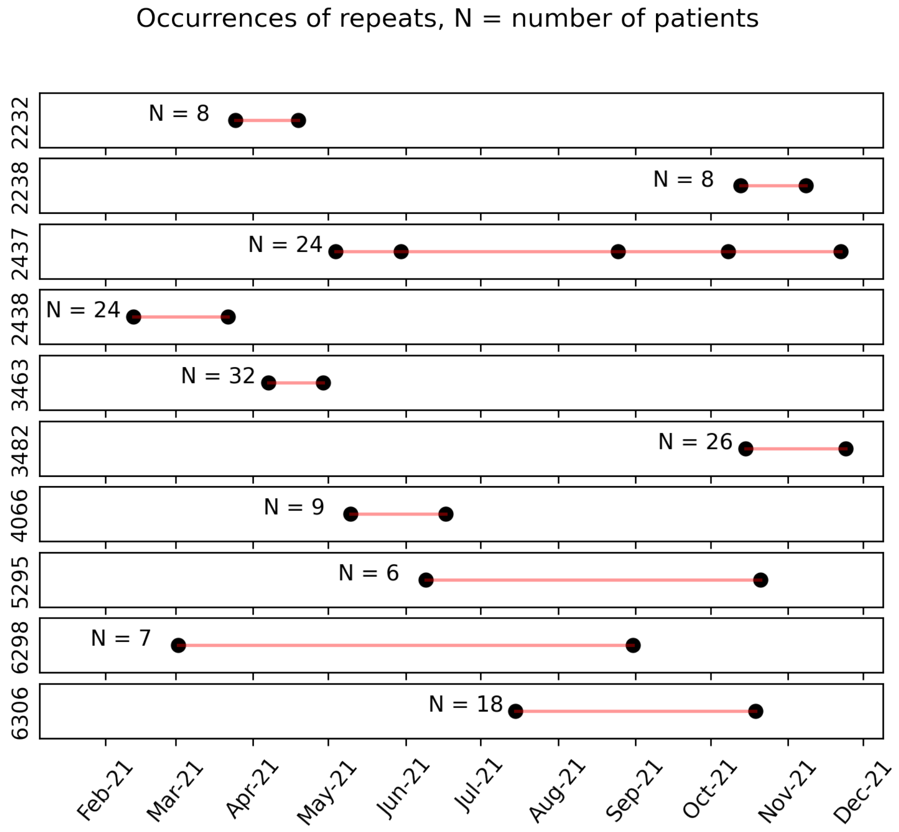


Figure 3: Selected examples of repeat requests separated by more than 21 days. Labels on the $y$ axis correspond to dataset IDs. Whilst some of these could be consistent with vaccination (ie, those with the longest interval), some repeats occur within around a month and so the explanation for these is uncertain.

Figure 3 shows further examples of repeat requests with an interval of at least 21 days. Some repeats are sufficiently spaced apart to be consistent with vaccination and/or booster doses, though such an explanation is insufficient for those separated by a month or so.

#### References

[1] John P Cole and Cuchlaine A M King. Quantitative geography: Techniques and theories in geography. Technical report, 1968.
